# Unsupervised phenotype clustering of non-ischemic dilated cardiomyopathy with AI-assisted T1 mapping cardiac MR

**DOI:** 10.64898/2026.07.10.26357725

**Authors:** Su A Noh, Hyun-Jin Kim, Kyeong Jin Park, Pan Ki Kim, Minjung Bak, Jiesuck Park, Hong-Mi Choi, Yeonyee E. Yoon, Goo-Yeong Cho, Byoung Wook Choi, Eun Ju Chun, In-Chang Hwang

## Abstract

**Aims:** Prognostic stratification and individual management are essential in the heterogeneous population of non-ischemic dilated cardiomyopathy (NIDCM). We applied unsupervised machine learning (ML) clustering in NIDCM cohorts, using semi-automated artificial intelligence (AI)-based cardiac magnetic resonance imaging (CMR) measurements with multimodal data to identify distinct phenotypes, characterize echocardiographic remodeling trajectories, and evaluate prognostic significance.

**Methods and results:** We analyzed 347 patients with NIDCM from two tertiary centers who underwent CMR and echocardiography at baseline, with follow-up echocardiography at a median 12 months. The cohort was randomly divided into derivation (n=242) and validation (n=105) sets using stratification by the composite outcome. Remodeling trajectories were evaluated using follow-up echocardiographic changes, and associations with outcomes were assessed by multivariable Cox regression adjusted for age and sex. Using eleven comprehensive clinical, laboratory, echocardiographic, and CMR-derived variables, partitioning around medoids clustering identified three phenotypes: (i) a younger, male-predominant preserved phenotype; (ii) a metabolic, fibrotic-remodeling phenotype; and (iii) an atrial fibrillation–predominant biventricular dysfunction phenotype. Cluster 1 showed the most favorable prognosis, whereas Cluster 3 had the highest risk of the composite outcome. Although LV reverse remodeling occurred across all clusters, Cluster 3 was characterized by attenuated LA reverse remodeling, suggesting persistent LA dysfunction.

**Conclusion:** Unsupervised ML-based clustering of NIDCM patients, integrating AI-derived CMR parameters with multimodal data, identified three clusters exhibiting distinct patterns in longitudinal echocardiographic trajectories and outcomes. This strategy may enable more individualized management in heterogeneous NIDCM.

## Introduction

Non-ischemic dilated cardiomyopathy (NIDCM) is characterized by left ventricular (LV) or biventricular dilatation and systolic dysfunction, in the absence of significant coronary artery disease or abnormal loading conditions.^1^ Although NIDCM is often regarded as a high-risk disease entity with the potential to progress to advanced heart failure, life-threatening arrhythmias, and cardiovascular death, its etiology and clinical course remain highly heterogeneous. This inherent variability underscores the need for refined prognostic stratification to individualize patient management.^2^

Current guidelines emphasize the role of multimodality imaging, including transthoracic echocardiography (TTE) and cardiac magnetic resonance imaging (CMR), for diagnosis and risk stratification of NIDCM. In particular, CMR enables detailed assessment of cardiac chamber remodeling and myocardial tissue characterization.^3^ Moreover, CMR–derived parameters—late gadolinium enhancement (LGE), native T1, and extracellular volume fraction (ECV)—have shown strong prognostic value beyond conventional TTE indices.^4–6^ Recent advances in artificial intelligence (AI)-based CMR quantification may enable more reproducible and efficient assessment of myocardial structure, tissue characteristics, and prognostic risk,^7^ thereby supporting broader clinical utilization of CMR-derived parameters.

The application of multi-parametric CMR data may contribute to a more refined understanding of disease heterogeneity in NIDCM. In parallel, machine learning (ML)–based clustering has emerged as a data-driven approach for identifying clinically meaningful phenotypes across various cardiovascular conditions.^8–11^ Such data-driven phenotyping can reveal latent disease subgroups that may not be captured by conventional clinical classification, and may therefore be particularly useful in heterogeneous cardiovascular diseases. However, evidence remains limited regarding the integration of CMR parameters for ML-based phenotyping in patients with NIDCM. Although previous studies have applied ML-based phenotyping to NIDCM or used CMR-derived tissue-characterization data for myocardial characterization,^12,13^ few have comprehensively integrated multiparametric CMR data into phenotyping and subsequently evaluated whether the derived phenotypic clusters differ in longitudinal cardiac functional trajectories and prognosis. This sequential approach is important because cluster-specific changes in cardiac function and clinical outcomes may clarify the clinical relevance of ML-derived phenotypes and their potential implications for treatment and management.

In this study, we applied semi-automated AI-based CMR quantification and unsupervised ML-based clustering in patients with NIDCM who underwent TTE and CMR. We aimed to identify distinct phenotypes, characterize their longitudinal remodeling trajectories, and evaluate their prognostic significance.

## Method

### Study Population

We conducted a retrospective cohort study of patients with NIDCM who underwent baseline CMR between January 2018 and December 2023 at two tertiary hospitals in South Korea: Seoul National University Bundang Hospital (SNUBH) and Hanyang University Guri Hospital (HYUMC-Guri).^14^ NIDCM was defined as LV dilation with systolic dysfunction (LVEF <50%) by the absence of the following conditions: (1) significant coronary artery disease (CAD) (≥50% luminal stenosis or prior myocardial infarction), (2) hypertensive heart disease, (3) tachycardia-induced or valvular cardiomyopathy, (4) infiltrative or inflammatory myocardial disease (sarcoidosis, amyloidosis, and myocarditis), (5) chemotherapy- or alcohol-induced cardiomyopathy, (6) burn-out hypertrophic cardiomyopathy, (7) LV non-compaction cardiomyopathy, or (8) arrhythmogenic right ventricular cardiomyopathy.^15^ Based on these criteria, a total of 347 patients were included in the final analysis.

The study protocol was approved under the Institutional Review Boards of both participating institutions (B-2406-905-104, GURI 2024-06-004), and informed consent was waived due to the retrospective design of the study and the use of anonymized data.

### Data collection

We retrospectively reviewed electronic medical records at the time of CMR study. TTE was performed using commercially available ultrasound systems and the parameters were measured according to the recommendations of the European Association of Cardiovascular Imaging.^16^ Follow-up echocardiography was reviewed at 6-18 months after the baseline study.

### CMR imaging acquisition and automated measurement of parameters

To minimize vendor- and site-related variability, we included only patients who underwent CMR using 3.0-T scanners from the same manufacturer and a standardized acquisition protocol at both institutions. CMR images were anonymized and all parameters were quantified using a semi-automated deep learning-based system (Myomics-T1 software, version 1.0.0, Phantomics Inc., Seoul, Republic of Korea), which provides automated segmentation and analysis. The development and validation of the deep learning algorithm have been previously described.^7,17,18^ Regions of interest were automatically delineated in the entire LV myocardium at basal, mid, and apical levels to calculate native T1 and ECV values. LGE extent was quantified using signal-intensity thresholds set at 5 and 6 standard deviations (SD) above remote myocardium, and both LGE mass and percentage were subsequently calculated. Quality control consisted of visual inspection and manual correction by two experienced CMR specialists who were blinded to clinical outcomes. Detailed CMR acquisition protocols are provided in the **Supplementary Materials**.

### Outcomes

The primary outcome was a composite of cardiovascular (CV) death and hospitalization for heart failure (HHF), adjudicated through a review of electronic medical records. Follow-up was calculated from the index date, defined as the date of baseline CMR, to the first occurrence of a clinical event or the last follow-up date, whichever came first. The secondary outcome was longitudinal change in echocardiographic parameters, including LV ejection fraction (LVEF), LV end-diastolic volume (LVEDV), LV end-systolic volume (LVESV), LA volume index (LAVI), and E/e’ ratio. For patients with multiple follow-up examinations, the study closest to 12 months after baseline was selected for analysis.

### Unsupervised machine learning-based clustering

The study population was randomly divided into derivation (70%; n = 242) and validation (30%; n = 105) cohorts using stratified sampling according to the occurrence of the primary outcome.^19^ This approach was chosen instead of a center-based split because of the substantial imbalance in sample size between participating centers, thereby maintaining adequate sample size and event distribution in both cohorts. The derivation cohort was used for cluster development, whereas the validation cohort was used to evaluate the reproducibility of cluster characteristics, echocardiographic trajectories, and clinical outcomes.

Candidate variables spanning clinical characteristics, laboratory values, echocardiographic parameters, and CMR parameters were selected based on clinical relevance and prior literature. Multicollinearity was assessed using correlation matrices and variance inflation factors, and principal component analysis was additionally performed to confirm minimal redundancy among the selected variables. Missing values were imputed (m = 30) using multivariate imputation by chained equations (MICE) with classification and regression trees (CART) after derivation–validation splitting to avoid information leakage.

Highly skewed continuous variables, such as N-terminal pro-B-type natriuretic peptide (NT-proBNP), were log-transformed, and ventricular volumes were indexed to body surface area (BSA). All continuous variables were then standardized to a mean of 0 and SD of 1, whereas binary variables were retained as 0/1 indicators.

The final clustering set comprised 11 variables: body-mass index (BMI), systolic blood pressure (SBP), estimated glomerular filtration rate (eGFR), atrial fibrillation (AF), log-transformed NT-proBNP (log NT-proBNP), TTE-derived LVGLS, BSA-indexed LVEDV (LVEDVi); CMR-derived LVEF, BSA-indexed right ventricular (RV) EDVi, native T1, and LGE extent (% by 5-SD threshold). Age and sex were not included in the clustering model; they were retained as covariates in the subsequent Cox regression analyses.

### Clustering algorithm and internal validation

To identify distinct clinical phenotypes, we performed partitioning around medoids (PAM) clustering based on the Euclidean distance matrix computed from the standardized variables. The optimal number of clusters was determined using quantitative metrics, clustering stability, and clinical interpretability. The relative contribution of each variable to the clustering solution was assessed using a random forest classifier with permutation importance. Internal validation was performed by assigning validation patients to the nearest derivation medoid and evaluating cluster reproducibility across cohorts.

### Statistical Analysis

Continuous variables, most of which were non-normally distributed (Shapiro–Wilk test), are summarized as median (interquartile range [IQR], 25th–75th percentile), and categorical variables as counts (%). Differences between the derivation and validation cohorts were assessed using the Mann–Whitney U test for continuous variables and the chi-square test or Fisher’s exact test for categorical variables. Differences across the three clusters were assessed using the Kruskal–Wallis test for continuous variables and the chi-square test for categorical variables.

Survival outcomes were analyzed using Kaplan–Meier curves and compared with log-rank tests. To assess the independent prognostic value of cluster membership, multivariable Cox proportional hazards models were constructed by adjusting for age and sex (Cluster 1 as the reference); hazard ratios (HR) and 95% confidence intervals (CI) were reported.

Temporal changes (Δ = follow-up – baseline) in echocardiographic parameters were analyzed. Within-cluster remodeling over time were assessed using the Wilcoxon signed-rank test. Differences in the magnitude of change across phenotypes were evaluated by analysis of covariance (ANCOVA) with the baseline value as a covariate, yielding the baseline-adjusted (least-square) mean change with 95% CI.

Imputation was performed in R (version 4.1.1; mice package), and all subsequent analyses were performed in Python (version 3.11; scipy, statsmodels, lifelines, and scikit-learn). A two-sided P < 0.05 was considered statistically significant.

## Results

### Clinical characteristics

Baseline characteristics of both cohorts are summarized in **Table 1**. Overall, the derivation and validation cohorts showed similar clinical profiles, including biventricular remodeling parameters, systolic function, and myocardial fibrosis burden. NT-proBNP levels and LAVI were higher in the validation cohort.

**Table 1.** Baseline Clinical and Multimodal Imaging Characteristics of the Study Cohort.

| Variable | Derivation (n=242) | Validation (n=105) | P |
| --- | --- | --- | --- |
| **Demographics and Physical Findings** |  |  |  |
| Age, years | 58.0 [45.3–70.2] | 56.6 [42.0–68.7] | 0.533 |
| Male, n (%) | 160 (66) | 74 (70) | 0.502 |
| Body-mass index, kg/m<sup>2</sup> | 24.3 [21.9–26.6] | 23.6 [21.6–25.9] | 0.126 |
| Systolic BP, mmHg | 120 [108–137] | 120 [107–136] | 0.506 |
| Diastolic BP, mmHg | 74.0 [66.0–83.0] | 74.0 [63.5–83.0] | 0.673 |
| Heart rate, beats/min | 81.0 [69.0–93.0] | 82.0 [67.0–92.0] | 0.615 |
| **Comorbidities, n (%)** |  |  |  |
| Hypertension | 56 (23) | 20 (19) | 0.480 |
| Diabetes mellitus | 52 (21) | 23 (22) | 1.000 |
| Dyslipidemia | 45 (19) | 24 (23) | 0.443 |
| Chronic kidney disease | 25 (10) | 7 (7) | 0.378 |
| COPD | 14 (6) | 6 (6) | 1.000 |
| Atrial fibrillation | 49 (20) | 21 (20) | 1.000 |
| Left bundle branch block | 27 (11) | 10 (10) | 0.792 |
| Coronary artery disease | 21 (9) | 8 (8) | 0.907 |
| Stroke | 16 (7) | 3 (3) | 0.203 |
| **Medication, n (%)** |  |  |  |
| RAS blockers | 235 (97) | 101 (96) | 0.740 |
| Beta blockers | 228 (94) | 98 (93) | 0.943 |
| MRA | 136 (56) | 74 (70) | 0.017 |
| SGLT2i | 57 (24) | 35 (33) | 0.078 |
| Ivabradine | 68 (28) | 29 (28) | 1.000 |
| **Laboratory findings** |  |  |  |
| Hemoglobin, g/dL | 14.2 [12.9–15.3] | 14.2 [12.5–15.5] | 0.745 |
| GFR, mL/min/1.73m<sup>2</sup> | 87.2 [66.3–100.3] | 90.7 [75.4–104.7] | 0.086 |
| NT-proBNP, pg/mL | 1694 [462–4558] | 2864 [1005–6255] | 0.012 |
| **Cardiac MRI** |  |  |  |
| LVEF, % | 28.0 [21.1–36.4] | 27.8 [18.8–37.0] | 0.822 |
| LVEDV, mL | 245 [202–296] | 241 [199–317] | 0.674 |
| LVESV, mL | 178 [123–231] | 175 [123–246] | 0.589 |
| LV mass, g | 194 [156–243] | 201 [159–256] | 0.372 |
| RVEF, % | 38.4 [26.4–47.4] | 38.5 [29.3–44.0] | 0.390 |
| RVEDV, mL | 134 [110–173] | 150 [110–185] | 0.243 |
| RVESV, mL | 82.2 [60.2–119.9] | 85.3 [64.1–131.6] | 0.276 |
| LGE presence, n (%) | 129 (53) | 66 (63) | 0.126 |
| LGE percent 5SD, % | 0.83 [0.00–2.99] | 1.42 [0.00–4.00] | 0.077 |
| LGE percent 6SD, % | 0.41 [0.00–1.92] | 0.79 [0.00–3.01] | 0.078 |
| Whole-myocardial Native T1, ms | 1326 [1294–1364] | 1330 [1305–1381] | 0.141 |
| Whole-myocardial ECV, % | 26.2 [24.0–29.0] | 27.3 [25.1–30.7] | 0.078 |
| Whole-myocardial T2, ms | 50.2 [47.9–52.4] | 51.4 [48.9–54.2] | 0.030 |
| **Echocardiography** |  |  |  |
| LVEDD, mm | 60.9 [56.0–64.0] | 62.5 [56.8–67.2] | 0.066 |
| LVESD, mm | 51.3 [46.0–56.8] | 52.0 [46.0–60.0] | 0.160 |
| LVEDV, mL | 159 [125–194] | 174 [129–215] | 0.114 |
| LVESV, mL | 112 [78–146] | 127 [86–164] | 0.060 |
| LVMI, g/m<sup>2</sup> | 132 [113–153] | 141 [113–158] | 0.288 |
| LVEF, % | 28.0 [22.7–36.4] | 24.8 [19.0–35.5] | 0.023 |
| LVGLS, % | 10.7 [7.8–13.9] | 10.5 [6.5–13.4] | 0.374 |
| LAVI, mL/m<sup>2</sup> | 50.6 [39.6–65.4] | 60.2 [41.8–72.9] | 0.037 |
| E/e' | 13.5 [9.9–19.4] | 14.5 [9.7–19.7] | 0.460 |
| TR Vmax, m/s | 2.40 [2.10–2.80] | 2.60 [2.25–3.00] | 0.055 |
| **Clinical outcomes, n (%)** |  |  |  |
| Follow-up duration (months) | 66.4 [41.9–89.8] | 58.1 [39.4–90.1] | 0.415 |
| Composite (CV death or HHF) | 106 (44) | 46 (44) | 1.000 |
| CV death | 7 (3) | 1 (1) | 0.443 |
| HHF | 104 (43) | 46 (44) | 0.979 |

Values are median [IQR] for continuous variables and n (%) for categorical variables. P values: Mann–Whitney U test (continuous) and chi-square or Fisher exact test (categorical). COPD, chronic obstructive pulmonary disease; RAS, renin-angiotensin system; LVEDD/LVESD, left ventricular end-diastolic/end-systolic dimension; RVEF, right ventricular ejection fraction; LVMI, LV mass index; TR, tricuspid regurgitation; CV, cardiovascular; HHF, hospitalization for heart failure.

### Clustering of the derivation cohort

The derivation cohort was stratified into three distinct phenotypes using PAM clustering and incorporating 11 multidimensional variables (**Figure1**, **Table 2**). Subsequent random forest-based variable importance analysis identified AF, CMR-derived LVEF, and log NT-proBNP as the major contributors to cluster discrimination (**Figure S1**). The unique clinical profiles of each cluster are visualized via radar plots in **Figure 2**, and baseline characteristics of the validation cohort are summarized in **Table S1**. Cluster 1 represented a younger, predominantly male phenotype with relatively favorable cardiac remodeling, whereas Cluster 2 was an elderly phenotype characterized by greater diabetic and cardiorenal burden and advanced LV remodeling. Cluster 3 exhibited an older phenotype with a high burden of AF accompanied by biventricular remodeling.

**Figure 1.**
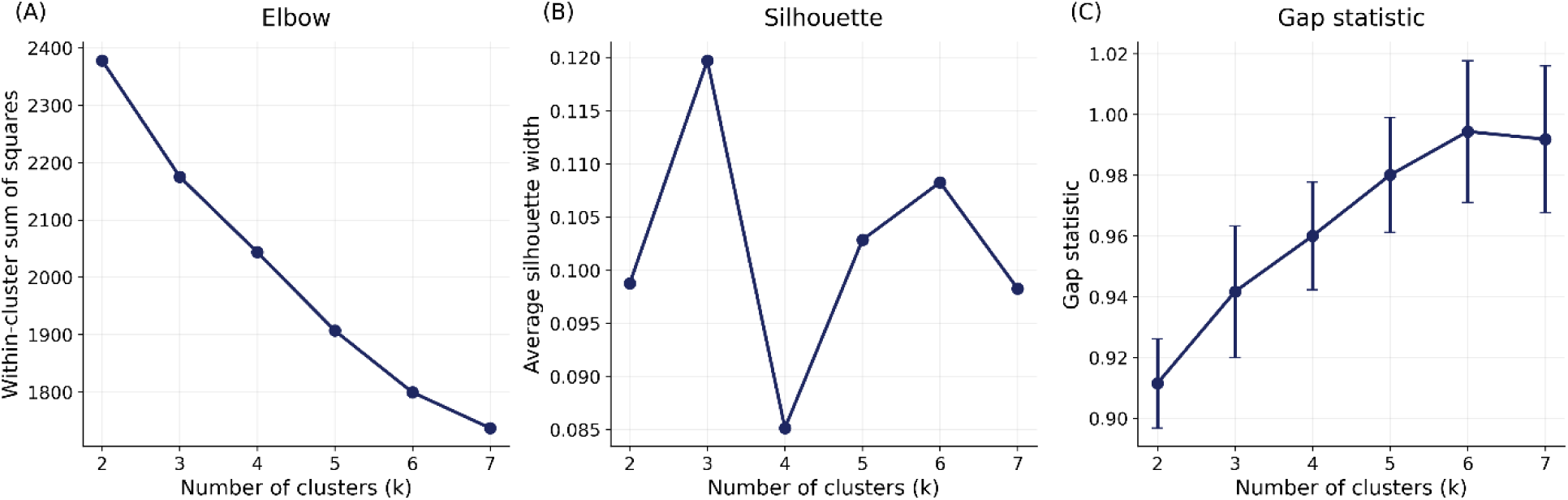
Selection of the optimal number of clusters. The optimal number of clusters (k) was determined using the elbow method (within-cluster sum of squares, WSS), silhouette score, and gap statistic.

**Figure 2.**
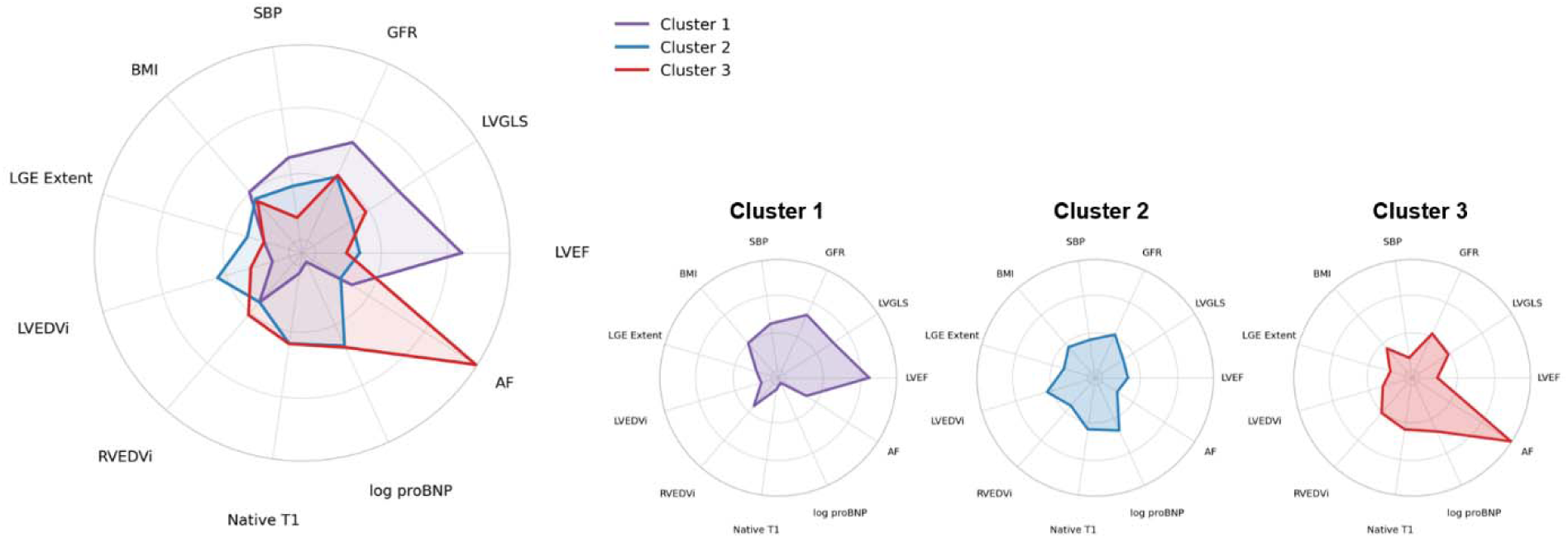
Radar plots of cluster phenotypes. Radar plots summarize clinical, echocardiographic, and CMR variables by cluster, including SBP, GFR, BMI, log NT-proBNP, AF prevalence, LVEF, LVGLS, LVEDVi, CMR-derived RVEDVi, native T1, and LGE extent (5-SD method). Continuous variables are presented as standardized cluster medians, whereas binary variables are presented as proportions.

**Table 2.** Clinical profile and associated clinical outcomes across clusters in derivation set.

| Variable | Cluster 1 (n=51) | Cluster 2 (n=145) | Cluster 3 (n=46) | P |
| --- | --- | --- | --- | --- |
| **Demographics and physical findings** |  |  |  |  |
| Age, years | 51.5 [36.5–60.2] | 59.3 [47.0–72.0] | 59.2 [47.8–71.4] | <0.001 |
| Male, n (%) | 37 (73) | 92 (63) | 31 (67) | 0.488 |
| Body-mass index, kg/m<sup>2</sup> | 24.7 [23.4–26.9] | 24.2 [21.6–26.4] | 24.0 [22.2–26.2] | 0.442 |
| Systolic BP, mmHg | 129 [118–144] | 120 [110–138] | 110 [103–123] | <0.001 |
| Diastolic BP, mmHg | 76.0 [69.0–82.5] | 74.0 [66.0–85.0] | 69.0 [62.2–75.8] | 0.043 |
| Heart rate, beats/min | 69.0 [63.0–83.0] | 83.0 [70.0–94.0] | 87.5 [71.5–99.8] | <0.001 |
| **Comorbidities, n (%)** |  |  |  |  |
| Hypertension | 11 (22) | 29 (20) | 16 (35) | 0.112 |
| Diabetes mellitus | 10 (20) | 35 (24) | 7 (15) | 0.410 |
| Dyslipidemia | 11 (22) | 26 (18) | 8 (17) | 0.825 |
| Chronic kidney disease | 3 (6) | 18 (12) | 4 (9) | 0.386 |
| COPD | 1 (2) | 8 (6) | 5 (11) | 0.168 |
| Atrial fibrillation | 4 (8) | 0 (0) | 45 (98) | <0.001 |
| Left bundle branch block | 4 (8) | 20 (14) | 3 (7) | 0.275 |
| Coronary artery disease | 2 (4) | 16 (11) | 3 (7) | 0.254 |
| Stroke | 2 (4) | 7 (5) | 7 (15) | 0.032 |
| **Medication, n (%)** |  |  |  |  |
| RAS blockers | 47 (92) | 142 (98) | 46 (100) | 0.046 |
| Beta blockers | 48 (94) | 135 (93) | 45 (98) | 0.489 |
| MRA | 23 (45) | 84 (58) | 29 (63) | 0.165 |
| SGLT2i | 10 (20) | 30 (21) | 17 (37) | 0.058 |
| Ivabradine | 10 (20) | 50 (34) | 8 (17) | 0.025 |
| **Laboratory findings** |  |  |  |  |
| Hemoglobin, g/dL | 14.4 [13.4–15.6] | 13.9 [12.6–15.1] | 14.6 [13.4–16.2] | 0.019 |
| GFR, mL/min/1.73m<sup>2</sup> | 99.3 [87.5–107.0] | 84.3 [62.5–98.6] | 85.0 [63.6–93.3] | <0.001 |
| NT-proBNP, pg/mL | 162 [56–549] | 2424 [946–5279] | 2513 [932–5785] | <0.001 |
| **Cardiac MR** |  |  |  |  |
| LVEF, % | 43.6 [34.5–50.1] | 25.9 [19.1–31.1] | 23.6 [20.0–31.3] | <0.001 |
| LVEDV, mL | 207 [170–230] | 267 [224–309] | 236 [196–295] | <0.001 |
| LVESV, mL | 116 [95–141] | 199 [155–247] | 173 [126–226] | <0.001 |
| LV mass, g | 160 [134–199] | 207 [176–249] | 198 [148–231] | <0.001 |
| RVEF, % | 44.5 [39.4–52.6] | 38.5 [24.4–47.8] | 32.0 [22.7–37.9] | <0.001 |
| RVEDV, mL | 131 [115–165] | 129 [106–174] | 147 [120–179] | 0.133 |
| RVESV, mL | 78.5 [53.1–92.1] | 80.6 [57.6–121.4] | 102 [79–126] | 0.003 |
| LGE presence, n (%) | 21 (41) | 88 (61) | 20 (43) | 0.019 |
| LGE percent 5SD, % | 0.00 [0.00–1.79] | 1.05 [0.00–3.69] | 0.00 [0.00–3.46] | 0.048 |
| LGE percent 6SD, % | 0.00 [0.00–1.33] | 0.64 [0.00–2.33] | 0.00 [0.00–2.69] | 0.062 |
| Whole-myocardial Native T1, ms | 1267 [1247–1313] | 1334 [1309–1378] | 1342 [1301–1368] | <0.001 |
| Whole-myocardial ECV, % | 25.5 [23.5–28.4] | 26.5 [24.1–29.0] | 26.2 [24.4–30.0] | 0.273 |
| Whole-myocardial T2, ms | 48.1 [47.2–50.0] | 50.5 [48.5–52.8] | 51.8 [49.8–53.3] | <0.001 |
| **Echocardiography** |  |  |  |  |
| LVEDD, mm | 58.5 [54.0–60.9] | 62.0 [58.0–65.1] | 60.0 [56.2–64.8] | <0.001 |
| LVESD, mm | 46.0 [41.2–51.0] | 52.3 [48.8–58.2] | 51.0 [47.0–55.9] | <0.001 |
| LVEDV, mL | 138 [114–160] | 168 [139–207] | 156 [124–190] | <0.001 |
| LVESV, mL | 77.0 [68.2–98.5] | 123 [96–160] | 111 [79–149] | <0.001 |
| LVMI, g/m<sup>2</sup> | 107 [96–128] | 143 [120–159] | 135 [117–156] | <0.001 |
| LVEF, % | 40.0 [33.9–46.5] | 26.1 [21.5–31.9] | 26.7 [22.6–34.2] | <0.001 |
| LVGLS, % | 13.5 [11.2–14.9] | 9.60 [7.38–12.60] | 10.9 [7.8–14.4] | <0.001 |
| LAVI, mL/m<sup>2</sup> | 35.9 [30.6–46.3] | 52.7 [42.2–67.3] | 61.0 [48.4–75.8] | <0.001 |
| E/e' | 8.85 [7.46–13.61] | 15.5 [10.9–20.6] | 13.2 [11.3–17.8] | <0.001 |
| TR Vmax, m/s | 2.20 [1.90–2.30] | 2.60 [2.20–3.00] | 2.50 [2.20–2.80] | <0.001 |
| **Clinical outcomes, n (%)** |  |  |  |  |
| Follow-up duration (months) | 67.2 [45.0–94.2] | 65.4 [42.1–82.9] | 72.9 [40.8–100.6] | 0.587 |
| Composite (CV death or HHF) | 12 (24) | 67 (46) | 27 (59) | 0.002 |
| CV death | 0 (0) | 4 (3) | 3 (7) | 0.158 |
| HHF | 12 (24) | 66 (46) | 26 (57) | 0.003 |

1 Abbreviations as in Table 1.

### Temporal changes in echocardiographic parameters according to clusters

During a median echocardiographic follow-up of 12.4 months (IQR 9.4–15.5), LV reverse remodeling was observed across all clusters in the derivation cohort, with significant improvements in LVEF and reductions in LV volumes (all p<0.001 for within-cluster changes) (**Figure 3, Table S2**). Despite this overall trend, LA reverse remodeling, as reflected by reductions in LAVI, appeared attenuated in Cluster 3. In the validation cohort, all three clusters demonstrated directionally consistent reverse remodeling patterns, except for the LAVI, with Cluster 3 again showing attenuated LAVI reduction compared with the other clusters. LVEF improved significantly in all clusters, whereas reductions in LV volumes, LAVI, and E/e′ within Cluster 1 did not reach statistical significance, likely due to the limited number of patients with follow-up echocardiographic data.

**Figure 3.**
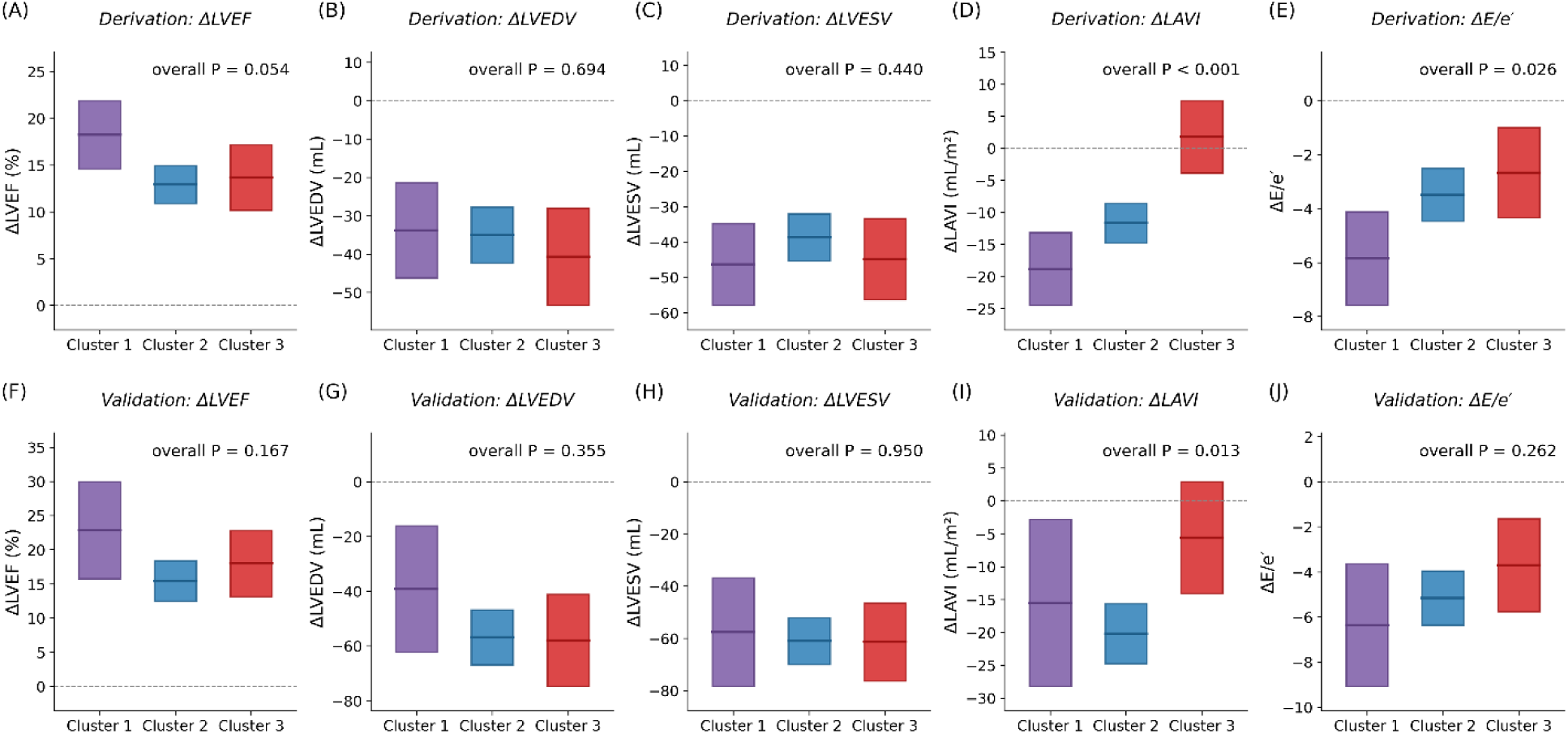
Baseline-adjusted echocardiographic changes by cluster. Floating bars show the baseline-adjusted (least-square) mean change (Δ = follow-up − baseline) with 95% confidence intervals, estimated using analysis of covariance (ANCOVA) with the baseline value as a covariate, stratified by cohort and echocardiographic parameter. Panels (A–E) depict the derivation cohort, and panels (F–J) the validation cohort, showing (from left to right) LVEF, LVEDV, LVESV, LAVI, and E/e′. The overall between-cluster P value from ANCOVA is presented in each panel. Positive Δ values indicate an increase from baseline to follow-up.

When baseline-adjusted changes were compared across clusters, LV reverse remodeling was broadly comparable across phenotypes in either cohort. In contrast, reductions in LAVI and E/e′ differed significantly across clusters in the derivation cohort, with the smallest reduction observed in Cluster 3 (overall P<0.001 for LAVI; overall P=0.026 for E/e′). Of these parameters, only LAVI reduction remained significantly different across clusters in the validation cohort (p=0.013).

### Adverse events according to clusters

After adjustment for age and sex, cluster-based differences in clinical outcomes were observed in the derivation cohort (**Figure 4**, **Table 3**). For the composite endpoint, event rates were largely driven by HHF, reflecting the low number of CV death events. A clear gradient in risk was evident across clusters, with the highest risk in Cluster 3 (adjusted HR 3.03, 95% CI 1.51–6.06; *p*=0.002), followed by Cluster 2 (adjusted HR, 2.33; 95% CI, 1.23-4.38; p=0.009), and the lowest risk in Cluster 1 (reference).

**Figure 4.**
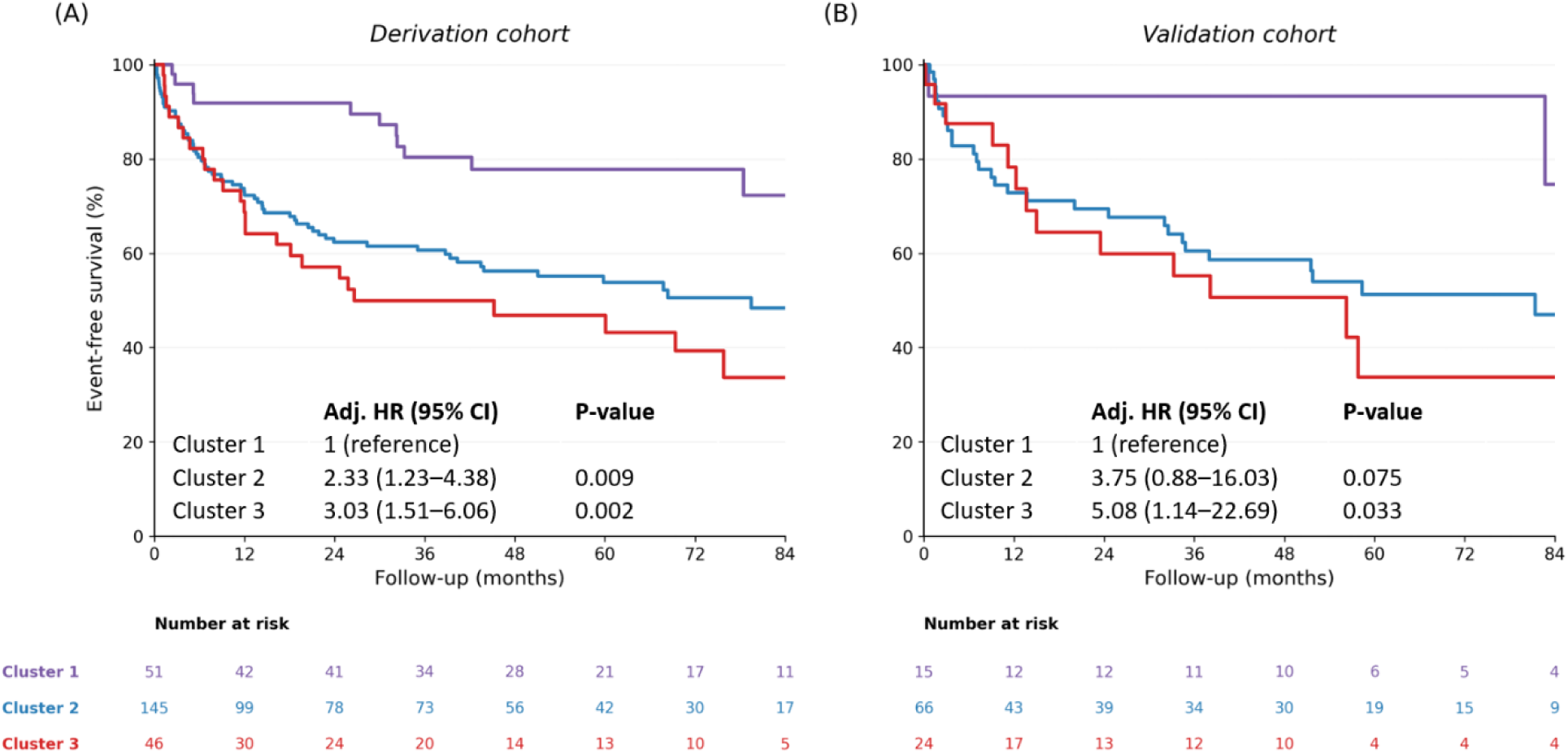
Survival analysis of composite outcomes according to clusters. Panels A and B show Kaplan–Meier curves for derivation and validation cohorts. Curves are stratified by cluster (1: purple, 2: blue, 3: red).

**Table 3.** Association between clusters and CV death or HHF.

| Variable | Derivation cohort |  | Validation cohort |  |
| --- | --- | --- | --- | --- |
|  | Adjusted HR (95% CI) | P | Adjusted HR (95% CI) | P |
| Age, per year | 1.01 (0.99–1.02) | 0.293 | 1.03 (1.01–1.06) | 0.004 |
| Sex (male) | 1.11 (0.73–1.67) | 0.627 | 1.32 (0.68–2.54) | 0.411 |
| Phenotype clusters |  |  |  |  |
| Cluster 1 | *Referenced* |  | *Referenced* |  |
| Cluster 2 | 2.33 (1.23–4.38) | 0.009 | 3.75 (0.88–16.03) | 0.075 |
| Cluster3 | 3.03 (1.51–6.06) | 0.002 | 5.08 (1.14–22.69) | 0.033 |

2 Multivariable Cox proportional-hazards model for the composite of CV death or HHF, including age, sex, and cluster membership 3 (Cluster 1 as reference).
Abbreviations: HR, hazards ratio; CI, confidence interval.

In the validation cohort, Cluster 3 remained significantly associated with higher risk (adjusted HR, 5.08; 95% CI, 1.14–22.69; p=0.033), while Cluster 2 showed a similar trend (adjusted HR, 3.75; 95% CI, 0.88–16.03; p=0.075). Results for HHF were generally consistent with those of the composite outcome and are presented in **Figure S2** and **Table S3**.

## Discussion

In this study, unsupervised ML clustering was applied to cohorts of patients with NIDCM who underwent multiparametric CMR. Three distinct phenotypes were identified: (i) a younger, male-predominant cluster with preserved cardiac function, (ii) an older cluster with diabetic–renal comorbidities, LBBB, and advanced LV fibrotic remodeling, and (iii) an older cluster with the greatest burden of AF, combined RV dysfunction. These phenotypes differed in remodeling patterns and clinical outcomes, highlighting the heterogeneity of NIDCM. To our knowledge, this is the first study to identify the distinct phenotypic clusters in patients with NIDCM by integrating multiparametric CMR data and to demonstrate cluster-specific differences in longitudinal remodeling trajectories and prognosis.

### Clinical heterogeneity of NIDCM

NIDCM is among the most prevalent forms of cardiomyopathy, characterized by diverse etiologies and a clinical spectrum, ranging from subclinical LV dysfunction to overt heart failure and sudden cardiac death.^4^ Although current guideline-based treatment recommendations still rely heavily on LVEF,^20,21^ LVEF alone incompletely captures the heterogeneous risk of cardiovascular events. Further, even among those with impaired LVEF, some do not respond adequately to optimal medical treatment and experience adverse cardiac remodeling and worse prognosis.^22,23^ These findings indicate the limitations of risk stratification and treatment guidance based solely on LVEF and highlight the needs to better understand the clinical and pathophysiological heterogeneity of NIDCM. In this context, CMR–derived parameters–including LGE, native T1, ECV–may provide important insights into heterogenous phenotypes of NIDCM through myocardial tissue characterization. Indeed, CMR-derived tissue characterization parameters have demonstrated incremental prognostic value beyond conventional functional indices, including LVEF, in patients with NIDCM.^24^ Nonetheless, each parameter has limitations when considered in isolation: LGE primarily detects focal fibrosis but may overlook diffuse interstitial changes, whereas native T1 and ECV reflect diffuse myocardial tissue changes but may be less sensitive to regional myocardial abnormalities.^25^ Consequently, a multiparametric approach that integrates imaging findings with clinical and laboratory data is desirable; however, a validated, standardized framework is lacking, often leaving physicians to rely on individual discretion.

### CMR-integrated clustering and myocardial tissue phenotyping in NIDCM

Emerging evidence indicates that unsupervised ML-based clustering aids in identifying previously unrecognized disease patterns by delineating discrete phenotypic clusters in various cardiovascular diseases.^8–11^ This approach may be particularly useful in diseases with substantial clinical and biological heterogeneity.^26^ Thus, ML-based phenotyping is highly relevant in NIDCM, in which etiology, pathophysiology, treatment response and prognosis vary widely across patients. Several previous studies have attempted to define distinct phenotypes among patients with DCM or heart failure. For example, Qiu et al. demonstrated a phenogroup of HF patients exhibiting metabolic, inflammatory, and body composition profiles indicative of impaired cardiorespiratory fitness,^9^ Gandin et al. identified two NIDCM subgroups based on baseline ECG variables, with distinct progression trajectories,^11^ and Verdonschot et al. described four DCM phenogroups with differing clinical outcomes using multimodal data.^27^ These findings support the concept that data-driven phenotyping may improve our understanding of disease heterogeneity in NIDCM.

Building on these insights, our clustering strategy was designed to integrate complementary clinical, laboratory, echocardiographic, and CMR domains. We explored multiple combinations of CMR parameters, including ECV, native T1, and LGE, to determine whether myocardial tissue characterization could improve phenotypic discrimination. This approach enabled identification of distinct phenotypes: Cluster 1 was characterized by a lower burden of myocardial fibrosis, reflected by low native T1 values and minimal LGE; Cluster 2 by advanced diffuse myocardial fibrosis, prevalent LGE and more pronounced LV dilation; and Cluster 3 by advanced diffuse fibrosis with more severe LA and RV remodeling. Notably, Cluster 3 showed less reversibility of cardiac remodeling and worse prognosis compared with Cluster 2, despite having less frequent replacement fibrosis on LGE. These findings highlight the unique value of CMR tissue characterization in distinguishing different stages or patterns of myocardial adverse remodeling in NIDCM.

Interestingly, although ECV is a well-established prognostic factor, its inclusion tended to reduce the reproducibility of the clustering solution in the present study.^6^ Among the combinations tested, native T1 and LGE yielded a clustering structure that was both stable and clinically interpretable. This may be because native T1 and LGE capture complementary aspects of myocardial tissue remodeling: native T1 reflects more diffuse interstitial abnormalities, whereas LGE represents more focal or advanced replacement fibrosis. Therefore, the combined use of native T1 and LGE may better represent the heterogeneous myocardial substrate of NIDCM.^28,29^

### Functional trajectories and outcomes across ML-derived phenotypes

Beyond myocardial tissue characteristics, the identified clusters were further differentiated by comorbidities and conventional echocardiographic parameters, underscoring the needs for comprehensive multidomain assessment in patients with NIDCM. These phenotypic distinctions were further supported by cluster-specific differences in longitudinal remodeling trajectories and prognosis. Notably, LV reverse remodeling occurred across all clusters, and baseline-adjusted changes in LV remodeling parameters were comparable among the three phenotypes. Given that the use of guideline-directed medical therapy (GDMT) did not differ significantly across clusters, these findings suggest that contemporary GDMT may contribute to attenuation of adverse LV remodeling across phenotypes. In contrast, LA reverse remodeling was attenuated in Cluster 3, which had the worst prognosis. Consistent with previous studies showing that concomitant improvement in both LA and LV function confers a more favorable prognosis than partial reverse remodeling,^22^ these findings suggest that impaired LA reverse remodeling may represent an important determinant of residual risk beyond LV functional recovery.

Importantly, the phenotypic clusters identified in the derivation cohort showed a consistent trend in the validation cohort, supporting the potential robustness of the clustering framework. Although further external validation in larger and more diverse population is warranted, these findings suggest that unsupervised ML-based clustering using multidomain clinical, echocardiographic, and CMR data can provide reproducible and clinically meaningful phenotyping, which may help refine personalized risk stratification and management in NIDCM.

### Utility of AI-assisted CMR parameters in NIDCM phenotyping

Another clinical relevance of the present study was the utilization of AI-assisted CMR parameters. Manual delineation of the endocardial and epicardial contours of the LV is necessary for the extraction of T1, ECV, and LGE values, but this process is labor-intensive and requires highly experienced experts to ensure accuracy and reproducibility. Additionally, it could introduce interobserver variability, limiting measurement consistency.^17,18^ To address these limitations, AI-assisted methods for quantifying CMR parameters have emerged as a promising solution, demonstrating robust validation in previous studies and improving both efficiency and reproducibility. Integrating AI-assisted CMR parameters into phenotypic clustering in NIDCM may overcome these limitations, reduce radiologist workload, and facilitate efficient and standardized analysis of CMR parameters.^17,30^

### Limitations

This study has several limitations. First, its retrospective design may have introduced selection and information biases. Second, generalizability may be limited because the study was conducted at only two tertiary centers in South Korea. In addition, the overall sample size was modest, and the validation cohort was particularly small (derivation, n = 242; validation, n = 105), with Cluster 1 including only 15 patients and 1 clinical event. The limited statistical power resulted in wide CI for HR estimates. Third, because the follow-up echocardiography was not performed according to prespecified schedules but at attending physicians’ discretion, some echocardiographic parameters (e.g., LVGLS) could not be analyzed due to missing follow-up data. Finally, biochemical analyses for pathophysiologic mechanisms or genetic tests or histopathology of myocardium were not included in the present study. Thus, we could not draw the pathobiologic background mechanisms for the phenotypic clusters. However, given the CMR tissue characterization showing distinct features by native T1 and LGE, we could provide more practical phenotypic clusters, which would more readily be accessible in real-world practice with better interpretability.

## Conclusions

Unsupervised ML-based clustering of patients with NIDCM, using AI-derived CMR parameters and multimodal data, identified clinically meaningful phenotypes with distinct patterns of cardiac reverse remodeling and prognosis. This approach highlights the potential for individualized management in heterogeneous NIDCM. Further studies with larger, externally validated cohorts are warranted to confirm and extend these findings.

## Data Availability

The data supporting the findings of this study are available from the corresponding author upon reasonable request.

## Notes

**Funding:** This work was supported by a grant from SNUBH (grant number: 06-2020-0130) and National Research Foundation of Korea (NRF) grant funded by the Korea government (Ministry of Science and ICT) (Grant No. RS-2024-00345490).

**Conflict of interest:** Pan Ki Kim and Byoung Wook Choi are founders of Phantomics, Inc. (Seoul, Korea), the company that supports the software used in this study. Phantomics, Inc. also funded the salaries of PKK and BWC but had no role in the study design, data collection, data analysis, decision to publish, or manuscript preparation. All other authors declare that they have no competing interests related to Phantomics, Inc., any other funders, or any institutions, including affiliations involving employment, consultancy, patents, products in development, or marketed products.

### Competing Interest Statement

Pan Ki Kim and Byoung Wook Choi are founders of Phantomics, Inc. (Seoul, Korea), the company that supports the software used in this study. Phantomics, Inc. also funded the salaries of PKK and BWC but had no role in the study design, data collection, data analysis, decision to publish, or manuscript preparation. All other authors declare that they have no competing interests related to Phantomics, Inc., any other funders, or any institutions, including affiliations involving employment, consultancy, patents, products in development, or marketed products.

### Author Declarations

The Institutional Review Boards of Seoul National University Bundang Hospital and Hanyang University Guri Hospital gave ethical approval for this work. The requirement for informed consent was waived because of the retrospective nature of the study.

