## Supplementary materials for "Unsupervised phenotype clustering of non-ischemic dilated cardiomyopathy with AI-assisted T1 mapping cardiac MR"

**Supplementary Methods: CMR Acquisition Protocol**

CMR was performed on a 3.0-T clinical scanner (Ingenia or Achieva; Philips Healthcare, Best, Netherlands) with a 16-channel phased array coil. Cine images using a steady-state free-precession sequence quantified LV and right ventricular (RV) volumes, mass, and ejection fraction. LGE imaging was obtained 10 minutes after intravenous administration of 0.15 mmol/kg gadobutrol using a phase-sensitive inversion-recovery sequence. Native T1-mapping images were acquired using a modified look-locker inversion-recovery 5(3)3 (MOLLI) sequence in three short-axis planes (basal, mid, and apical of the LV), while post-contrast T1 mapping was performed 15 minutes after contrast administration using a 4(1)3(1)2 sequence along the same three planes. Typical imaging parameters for T1 mapping were as follows: flip angle, 35°; field of view, 300 × 300 mm²; matrix size, 192 × 166; slice thickness, 8 mm; and acquisition window, 160–200 ms. All mapping images were acquired during a single breath-hold at end-diastole. ECV was calculated as ECV = (1–hematocrit) × [△R1_myocardium_]/[△R1_blood_], with hematocrit measured at the time of CMR imaging, where ΔR1 is the change in relaxivity and determined using the following equation: △R1_myocardium_ = R1_post-myocardium_ – R1_pre-myocardium_, and △R1_blood=_ R1_post-blood_ – R1_pre-blood_.(1)

Quantitative tissue parameters were analyzed using an AI-based segmentation and mapping platform (Myomics, Seoul, Republic of Korea). Detailed information, including development and validation, on the AI-algorithm is provided in previous studies.(2-4) The algorithm automatically delineated endocardial and epicardial contours on basal, mid, and apical short-axis slices to compute native T1, post-T1, and ECV values for the entire LV myocardium (whole-myocardial) and the mid-septal region. T1 and ECV maps were generated from the original MOLLI DICOM images exported to the post-processing software, rather than from inline reconstructions. To minimize the partial volume effect at the endocardial and epicardial borders, the algorithm applied an automated 10% internal erosion to the contours, ensuring that T1 and ECV values were sampled exclusively from the mid-wall myocardium. The platform includes integrated motion correction (MOCO) and rigid/non-rigid co-registration between pre- and post-contrast T1 maps to ensure accurate ECV calculation. The entire myocardium was included in T1 and ECV measurements—LGE areas were not excluded—reflecting the total burden of both diffuse and focal fibrosis. Quality control included visual inspection and manual correction by two experienced CMR specialists who were blinded to clinical outcomes. Minor manual adjustments to the endocardial or epicardial contours were required in 5.8% of cases, primarily to refine boundary definitions. Slices with significant artifacts or irreversible co-registration errors were excluded from the final analysis; however, the three-slice (basal, mid, apical) acquisition ensured that a representative global value could be maintained even when an individual slice was suboptimal. LGE quantification was performed using the same AI-assisted segmentation platform for consistency. A reference region of interest was manually placed in a remote, visually normal (dark) myocardial area without focal enhancement or artifacts. LGE extent was then quantified automatically using signal-intensity thresholds of 5 SD (standard deviation) and 6 SD above the mean signal intensity of the remote reference region, applied to phase-sensitive inversion-recovery (PSIR) reconstruction images, and LGE mass and LGE percentage were calculated.

**Figure S1. Variable importance for cluster discrimination**

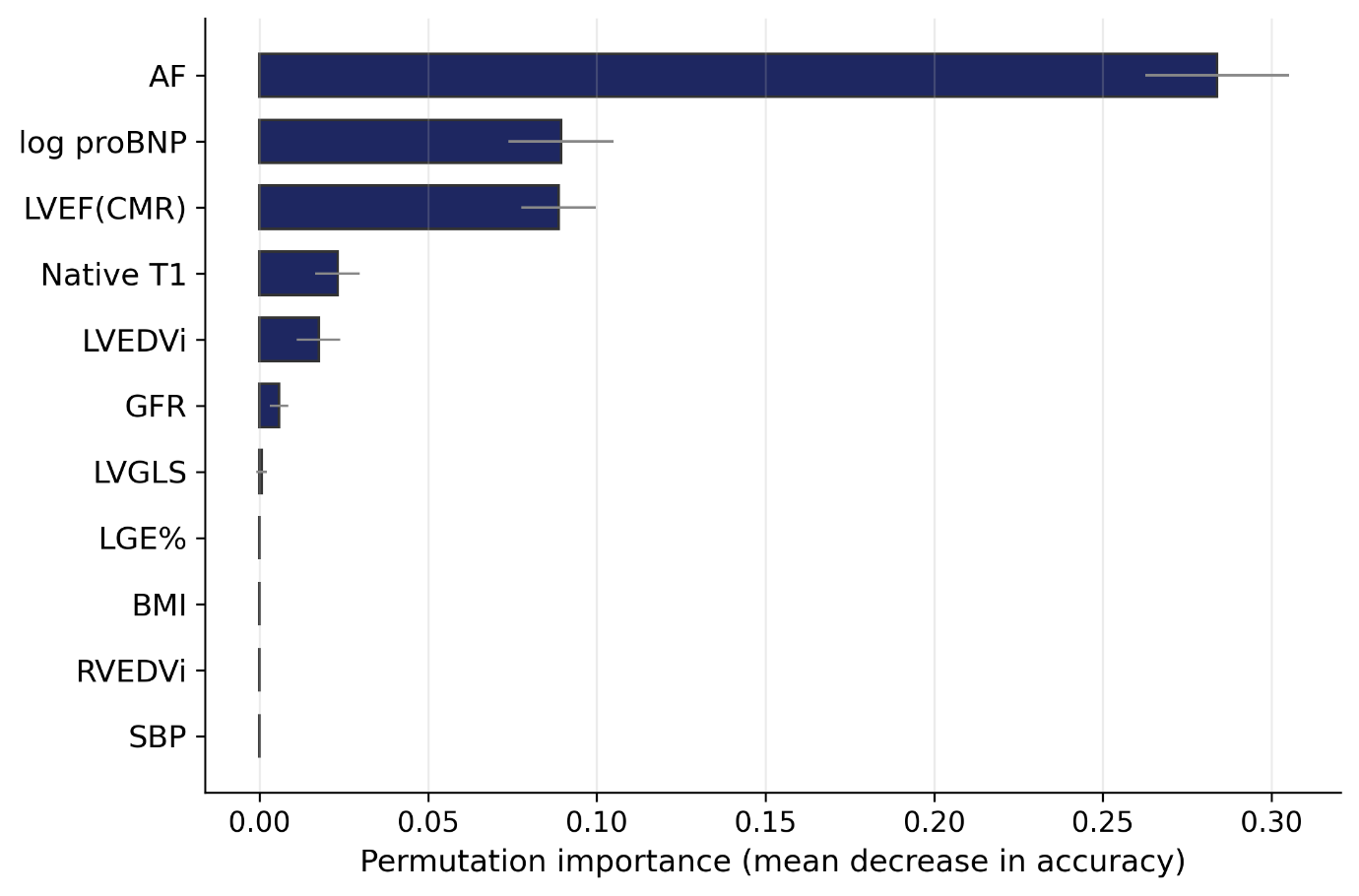

**Table S1. Clinical profile and associated clinical outcomes across clusters in validation set**

| **Variable** | **Cluster 1 (n=15)** | **Cluster 2 (n=66)** | **Cluster 3 (n=24)** | **P** |
| --- | --- | --- | --- | --- |
| **Demographics and Physical Findings** | | | | |
| Age, years | 45.9 [39.9–57.8] | 60.2 [45.1–71.2] | 55.8 [41.9–68.8] | 0.047 |
| Male, n (%) | 14 (93) | 41 (62) | 19 (79) | 0.033 |
| Body-mass index, kg/m² | 24.4 [23.5–25.7] | 22.7 [19.7–25.2] | 25.1 [23.4–26.6] | 0.004 |
| Systolic BP, mmHg | 135 [118–139] | 121 [106–137] | 110 [104–118] | 0.003 |
| Diastolic BP, mmHg | 80.0 [74.5–83.5] | 75.0 [65.5–83.8] | 68.0 [62.0–74.0] | 0.046 |
| Heart rate, beats/min | 79.0 [67.0–86.5] | 81.5 [66.2–92.0] | 87.5 [70.0–94.2] | 0.279 |
| **Comorbidities, n (%)** | | | | |
| Hypertension | 2 (13) | 11 (17) | 7 (29) | 0.341 |
| Diabetes mellitus | 1 (7) | 17 (26) | 5 (21) | 0.269 |
| Dyslipidemia | 0 (0) | 15 (23) | 9 (38) | 0.025 |
| Chronic kidney disease | 2 (13) | 5 (8) | 0 (0) | 0.238 |
| COPD | 2 (13) | 3 (5) | 1 (4) | 0.389 |
| Atrial fibrillation | 0 (0) | 0 (0) | 21 (88) | <0.001 |
| Left bundle branch block | 0 (0) | 8 (12) | 2 (8) | 0.344 |
| Coronary artery disease | 0 (0) | 6 (9) | 2 (8) | 0.482 |
| Stroke | 0 (0) | 3 (5) | 0 (0) | 0.402 |
| **Medication, n (%)** | | | | |
| RAS blockers | 14 (93) | 65 (98) | 22 (92) | 0.269 |
| Beta blockers | 13 (87) | 61 (92) | 24 (100) | 0.238 |
| MRA | 9 (60) | 49 (74) | 16 (67) | 0.494 |
| SGLT2i | 3 (20) | 23 (35) | 9 (38) | 0.483 |
| Ivabradine | 3 (20) | 26 (39) | 0 (0) | <0.001 |
| **Laboratory findings** | | | | |
| Hemoglobin, g/dL | 15.0 [14.2–15.7] | 13.9 [12.1–15.4] | 14.4 [12.8–15.7] | 0.109 |
| GFR, mL/min/1.73m² | 96.6 [87.4–107.2] | 88.9 [72.7–103.6] | 86.4 [74.0–104.5] | 0.179 |
| NT-proBNP, pg/mL | 300 [58–550] | 3507 [1424–6247] | 3840 [1054–6767] | 0.001 |
| **Cardiac MRI** | | | | |
| LVEF, % | 44.2 [40.4–48.4] | 24.6 [17.8–33.8] | 27.3 [23.0–34.2] | <0.001 |
| LVEDV, mL | 182 [171–228] | 259 [214–327] | 244 [202–287] | 0.003 |
| LVESV, mL | 102 [91–130] | 196 [145–267] | 174 [145–215] | <0.001 |
| LV mass, g | 156 [150–197] | 218 [169–280] | 191 [161–225] | 0.020 |
| RVEF, % | 44.0 [40.9–46.2] | 37.4 [25.7–43.6] | 38.9 [29.2–42.0] | 0.009 |
| RVEDV, mL | 161 [139–177] | 136 [103–187] | 157 [135–182] | 0.394 |
| RVESV, mL | 84.5 [80.2–99.3] | 80.1 [58.3–143.2] | 94.5 [81.7–137.5] | 0.565 |
| LGE presence, n (%) | 5 (33) | 48 (73) | 13 (54) | 0.010 |
| LGE percent 5SD, % | 0.00 [0.00–0.38] | 1.58 [0.00–4.97] | 2.79 [0.00–5.47] | 0.007 |
| LGE percent 6SD, % | 0.00 [0.00–0.14] | 0.97 [0.00–3.11] | 1.91 [0.00–3.61] | 0.007 |
| Whole-myocardial Native T1, ms | 1255 [1238–1294] | 1348 [1314–1398] | 1327 [1313–1344] | <0.001 |
| Whole-myocardial ECV, % | 24.1 [21.9–25.8] | 27.8 [25.5–31.3] | 28.2 [26.1–30.3] | 0.017 |
| Whole-myocardial T2, ms | 47.7 [46.6–48.6] | 51.8 [49.4–54.2] | 52.0 [50.2–54.6] | 0.029 |
| **Echocardiography** | | | | |
| LVEDD, mm | 56.0 [54.0–60.0] | 64.0 [59.0–69.4] | 62.0 [56.0–65.0] | <0.001 |
| LVESD, mm | 43.0 [39.5–47.0] | 56.0 [49.2–61.8] | 51.0 [45.9–56.5] | <0.001 |
| LVEDV, mL | 116 [98–150] | 184 [144–229] | 164 [109–208] | 0.001 |
| LVESV, mL | 66.0 [49.5–86.5] | 140 [100–172] | 104 [85–146] | <0.001 |
| LVMI, g/m² | 105 [80–121] | 146 [130–175] | 139 [102–148] | <0.001 |
| LVEF, % | 40.5 [30.9–46.1] | 22.0 [18.0–29.2] | 30.6 [21.3–34.4] | <0.001 |
| LVGLS, % | 13.2 [9.9–13.5] | 9.20 [5.30–13.00] | 11.8 [9.4–13.7] | 0.041 |
| LAVI, mL/m² | 36.1 [27.6–37.1] | 61.9 [46.4–77.7] | 63.8 [51.4–72.9] | <0.001 |
| E/e′ | 8.14 [7.25–10.22] | 16.3 [12.1–21.9] | 13.9 [9.7–18.4] | <0.001 |
| TR Vmax, m/s | 2.30 [2.15–2.40] | 2.60 [2.40–3.00] | 2.60 [2.30–2.95] | 0.079 |
| **Clinical outcomes, n (%)** | | | | |
| Follow-up duration (months) | 52.2 [41.0–88.1] | 62.5 [43.1–86.1] | 50.5 [35.9–97.6] | 0.924 |
| Composite (CV death or HHF) | 2 (13) | 30 (45) | 14 (58) | 0.020 |
| CV death | 0 (0) | 1 (2) | 0 (0) | 0.742 |
| Hospitalization for heart failure | 2 (13) | 30 (45) | 14 (58) | 0.020 |

Values are median [IQR] for continuous variables and n (%) for categorical variables. P values: Mann–Whitney U test (continuous) and chi-square or Fisher exact test (categorical). COPD, chronic obstructive pulmonary disease; RAS, renin-angiotensin system; LVEDD/LVESD, left ventricular end-diastolic/end-systolic dimension; RVEF, right ventricular ejection fraction; LVMI, LV mass index; TR, tricuspid regurgitation; CV, cardiovascular; HHF, hospitalization for heart failure.

**Table S2. Differential longitudinal LV remodeling pattern according to clusters in derivation set**

| **Variable** |  | **Cluster 1 (n=51)** | **Cluster 2 (n=145)** | **Cluster 3 (n=46)** | **P** |
| --- | --- | --- | --- | --- | --- |
| LVEF, % | Baseline | 39.5 [34.1–46.1] | 26.1 [21.4–31.6] | 26.6 [22.2–34.0] |  |
|  | Follow-up | 49.0 [44.5–54.5] | 42.2 [32.0–52.1] | 41.9 [34.0–53.0] |  |
|  | Adjusted mean change (95% CI) | +18.2 (+14.6 to +21.9) | +12.9 (+10.9 to +14.9) | +13.7 (+10.2 to +17.2) | 0.054 |
| LVEDV, mL | Baseline | 140 [114–161] | 168 [139–207] | 153 [122–192] |  |
|  | Follow-up | 112 [91–143] | 126 [96–161] | 113 [94–133] |  |
|  | Adjusted mean change (95% CI) | -33.8 (-46.2 to -21.4) | -35.1 (-42.3 to -27.8) | -40.7 (-53.4 to -28.1) | 0.694 |
| LVESV, mL | Baseline | 77.0 [68.0–99.5] | 124 [98–160] | 111 [78–150] |  |
|  | Follow-up | 54.7 [44.3–71.9] | 71.5 [46.0–102.4] | 63.5 [46.1–77.4] |  |
|  | Adjusted mean change (95% CI) | -46.4 (-57.9 to -34.8) | -38.7 (-45.3 to -32.0) | -44.9 (-56.4 to -33.4) | 0.440 |
| LAVI, mL/m² | Baseline | 35.9 [30.5–44.2] | 52.0 [41.3–67.3] | 63.9 [47.9–76.8] |  |
|  | Follow-up | 31.2 [25.1–39.2] | 39.2 [31.6–52.3] | 55.7 [39.6–78.8] |  |
|  | Adjusted mean change (95% CI) | -18.8 (-24.5 to -13.2) | -11.7 (-14.8 to -8.6) | +1.8 (-3.9 to +7.4) | <0.001 |
| E/e′ | Baseline | 9.00 [7.45–13.51] | 15.7 [10.9–20.5] | 13.2 [11.2–17.3] |  |
|  | Follow-up | 8.66 [6.53–10.43] | 10.7 [8.2–14.8] | 11.6 [8.3–15.6] |  |
|  | Adjusted mean change (95% CI) | -5.9 (-7.6 to -4.1) | -3.5 (-4.5 to -2.5) | -2.7 (-4.3 to -1.0) | 0.026 |

Data are presented as median [IQR]. Adjusted mean change represents the baseline-adjusted least-squares mean change (95% CI) estimated using analysis of covariance (change ~ cluster + baseline) based on complete-case paired data. P values represent the overall effect of cluster in the analysis of covariance model.

Abbreviations: LVEF, left ventricular ejection fraction; LVEDV/LVESV, left ventricular end-diastolic/end-systolic volume; LAVI, left atrial volume index; CI, confidence interval.

**Table S3. Differential longitudinal LV remodeling pattern according to clusters in validation set**

| **Variable** |  | **Cluster 1 (n=15)** | **Cluster 2 (n=66)** | **Cluster 3 (n=24)** | **P** |
| --- | --- | --- | --- | --- | --- |
| LVEF, % | Baseline | 38.3 [26.3–45.1] | 22.0 [18.0–29.2] | 28.9 [20.4–33.7] |  |
|  | Follow-up | 52.8 [43.7–58.8] | 44.4 [34.6–48.6] | 45.8 [35.4–54.3] |  |
|  | Adjusted mean change (95% CI) | +22.9 (+15.8 to +30.0) | +15.4 (+12.5 to +18.4) | +18.0 (+13.1 to +22.8) | 0.167 |
| LVEDV, mL | Baseline | 134 [98–161] | 185 [144–229] | 164 [109–218] |  |
|  | Follow-up | 128 [96–148] | 122 [92–161] | 103 [80–148] |  |
|  | Adjusted mean change (95% CI) | -39.2 (-62.3 to -16.1) | -56.8 (-66.9 to -46.8) | -58.0 (-74.7 to -41.2) | 0.355 |
| LVESV, mL | Baseline | 80.5 [52.7–97.8] | 141 [100–180] | 116 [83–158] |  |
|  | Follow-up | 62.0 [43.5–73.0] | 67.0 [48.0–103.0] | 58.2 [36.0–92.0] |  |
|  | Adjusted mean change (95% CI) | -57.6 (-78.3 to -36.8) | -61.0 (-70.0 to -52.1) | -61.3 (-76.2 to -46.5) | 0.950 |
| LAVI, mL/m² | Baseline | 33.8 [28.5–36.8] | 64.4 [47.4–79.4] | 63.8 [51.8–70.6] |  |
|  | Follow-up | 31.3 [28.8–36.1] | 37.0 [33.4–47.2] | 58.1 [34.4–74.8] |  |
|  | Adjusted mean change (95% CI) | -15.5 (-28.2 to -2.8) | -20.2 (-24.7 to -15.6) | -5.6 (-14.1 to +2.9) | 0.013 |
| E/e′ | Baseline | 8.09 [7.52–10.16] | 15.8 [11.5–23.0] | 13.9 [9.8–18.0] |  |
|  | Follow-up | 8.01 [6.92–9.42] | 9.88 [8.29–13.21] | 9.18 [8.31–14.14] |  |
|  | Adjusted mean change (95% CI) | -6.4 (-9.1 to -3.6) | -5.2 (-6.4 to -4.0) | -3.7 (-5.8 to -1.6) | 0.262 |

Data are presented as median [IQR]. Adjusted mean change represents the baseline-adjusted least-squares mean change (95% CI) estimated using analysis of covariance (change ~ cluster + baseline) based on complete-case paired data. P values represent the overall effect of cluster in the analysis of covariance model.

Abbreviations: LVEF, left ventricular ejection fraction; LVEDV/LVESV, left ventricular end-diastolic/end-systolic volume; LAVI, left atrial volume index; CI, confidence interval.

**Figure S2. Survival analysis of heart failure hospitalization according to clusters.**

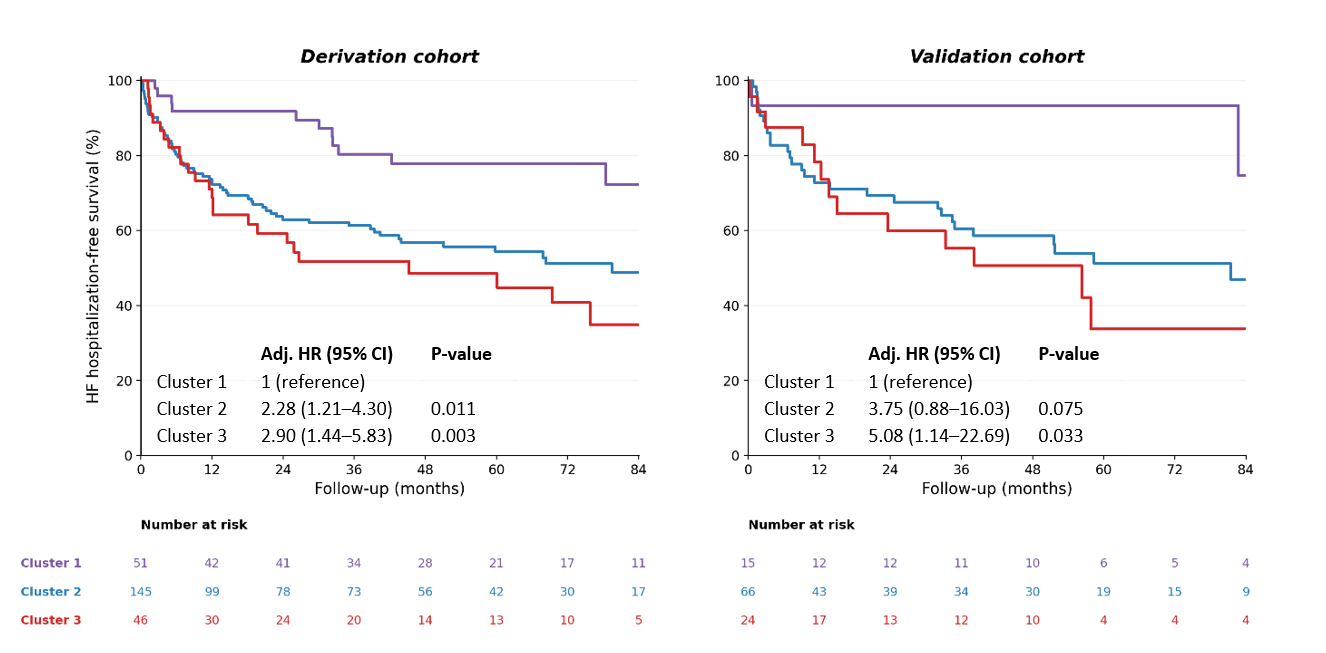

Panels A and B show Kaplan–Meier curves for derivation and validation cohorts. Curves are stratified by cluster (1: purple, 2: blue, 3: red).

**Table S4. Association Between Clusters and HHF**

|  | **Derivation cohort** | |  | **Validation cohort** | |
| --- | --- | --- | --- | --- | --- |
| **Variable** | **Adjusted HR (95% CI)** | **P** |  | **Adjusted HR (95% CI)** | **P** |
| Age, per year | 1.01 (0.99–1.02) | 0.258 |  | 1.03 (1.01–1.06) | 0.004 |
| Sex (male) | 1.18 (0.77–1.79) | 0.444 |  | 1.32 (0.68–2.54) | 0.411 |
| Phenotype clusters |  |  |  |  |  |
| Cluster 1 | *Referenced* |  |  | *Referenced* |  |
| Cluster 2 | 2.28 (1.21–4.30) | 0.011 |  | 3.75 (0.88–16.03) | 0.075 |
| Cluster3 | 2.90 (1.44–5.83) | 0.003 |  | 5.08 (1.14–22.69) | 0.033 |
